# Omicron mutations enhance infectivity and reduce antibody neutralization of SARS-CoV-2 virus-like particles

**DOI:** 10.1101/2021.12.20.21268048

**Authors:** Abdullah M. Syed, Alison Ciling, Mir M. Khalid, Bharath Sreekumar, Pei-Yi Chen, G. Renuka Kumar, Ines Silva, Bilal Milbes, Noah Kojima, Victoria Hess, Maria Shacreaw, Lauren Lopez, Matthew Brobeck, Fred Turner, Lee Spraggon, Taha Y. Taha, Takako Tabata, Irene P. Chen, Melanie Ott, Jennifer A. Doudna

## Abstract

The Omicron SARS-CoV-2 virus contains extensive sequence changes relative to the earlier arising B.1, B.1.1 and Delta SARS-CoV-2 variants that have unknown effects on viral infectivity and response to existing vaccines. Using SARS-CoV-2 virus-like particles (SC2-VLPs), we examined mutations in all four structural proteins and found that Omicron showed 3-fold higher capsid assembly and cell entry relative to Delta, a property conferred by S and N protein mutations. Thirty-eight antisera samples from individuals vaccinated with Pfizer/BioNTech, Moderna, Johnson & Johnson vaccines and convalescent sera from unvaccinated COVID-19 survivors had 15-fold lower efficacy to prevent cell transduction by VLPs containing the Omicron mutations relative to the ancestral B.1 spike protein. A third dose of Pfizer vaccine elicited substantially higher neutralization titers against Omicron, resulting in detectable neutralizing antibodies in 8 out of 8 subjects compared to 1 out of 8 pre-boost. Furthermore, the monoclonal antibody therapeutics Casirivimab and Imdevimab had robust neutralization activity against B.1, B.1.1 or Delta VLPs but no detectable neutralization of Omicron VLPs. Our results suggest that Omicron is more efficient at assembly and cell entry compared to Delta, and the antibody response triggered by existing vaccines or previous infection, at least prior to boost, will have limited ability to neutralize Omicron. In addition, some currently available monoclonal antibodies will not be useful in treating Omicron-infected patients.

**One-Sentence Summary:** Omicron SARS-CoV-2 virus-like particles have enhanced infectivity that is only weakly neutralized by vaccination without boost or prior infection, or antibody therapeutics.

---

Understanding the molecular determinants of SARS-CoV-2 viral fitness is central to effective vaccine and therapeutic development. The emergence of viral variants including Delta and Omicron underscores the need to assess both infectivity and antibody neutralization, but biosafety level 3 (BSL-3) handling requirements slow the pace of research on the intact SARS-CoV-2 virus. Although vesicular stomatitis virus (VSV) and lentiviruses pseudotyped with the SARS-CoV-2 spike (S) protein enable evaluation of S-mediated cell binding and entry via the ACE2 and TMPRSS2 receptors, they cannot determine effects of mutations outside the S gene (*1, 2*). To address these challenges, we developed SARS-CoV-2 virus-like particles (SC2-VLPs) comprising the S, N, M and E structural proteins and a packaging signal-containing messenger RNA that together form RNA-loaded capsids capable of spike-dependent cell transduction (*3*). This system faithfully reflects the impact of mutations in structural proteins that are observed in live-virus infections, enabling rapid testing of SARS-CoV-2 structural gene variants for their impact on both infection efficiency and antibody or anti serum neutralization.

Using the SC2-VLP system, we first generated a set of plasmid constructs encoding the S, N, M and E structural proteins derived from the B.1, B.1.1, Delta and Omicron SARS-CoV-2 viral variants (Fig. 1A). We generated SC2-VLPs by co-transfecting packaging cells (HEK293T cells) with three plasmids encoding these structural proteins and a fourth plasmid encoding luciferase mRNA linked to a SARS-CoV-2 packaging signal (*3*). Particles secreted from these packaging cells were filtered and incubated with receiver 293T cells stably co-expressing ACE2 and TMPRSS2 (Fig. 1B). To compare the effects of the different structural gene variants on infectivity, we used the structural genes from SARS-CoV-2 B.1 as the point of reference from which to vary each structural gene individually since it is ancestral to all currently circulating variants. We first tested the effects on infectivity of variation of the S gene in VLPs that otherwise contained B.1 gene versions. We found that relative to S-B.1 (identical to S-B.1.1), the Delta S variant produced VLPs that were only 20% as infectious (Fig. 1C). In contrast, the Omicron S gene in the context of the B.1 background generated VLPs that were at least as infectious as the ancestral S-B.1 (Fig. 1C). We also generated S variants containing Omicron mutations outside the receptor binding domain (RBD) but containing only mutations within the receptor binding domain (RBD) previously shown to inhibit binding by Class 1 (417N, 496S, 498R, 501Y) or Class 3 (440K, 446S, 496S, 498R) antibodies (*4*). These Omicron variants displayed moderately enhanced infectivity of 1.8- and 1.5-fold (S-OmC1, S-OmC3, Fig. 1C, Supplementary Table S1). These results are in contrast to observations of pseudotyped lentiviral particles which show increased entry for S-Delta and reduced entry for S-Omicron (Supplementary Fig. S1). These results suggest that viral genetic context influences S gene effects on the ability of viral particles to transduce cells, and also that some S gene mutations such as those in Omicron may dominate cell infectivity outcomes.

**Fig. 1.**
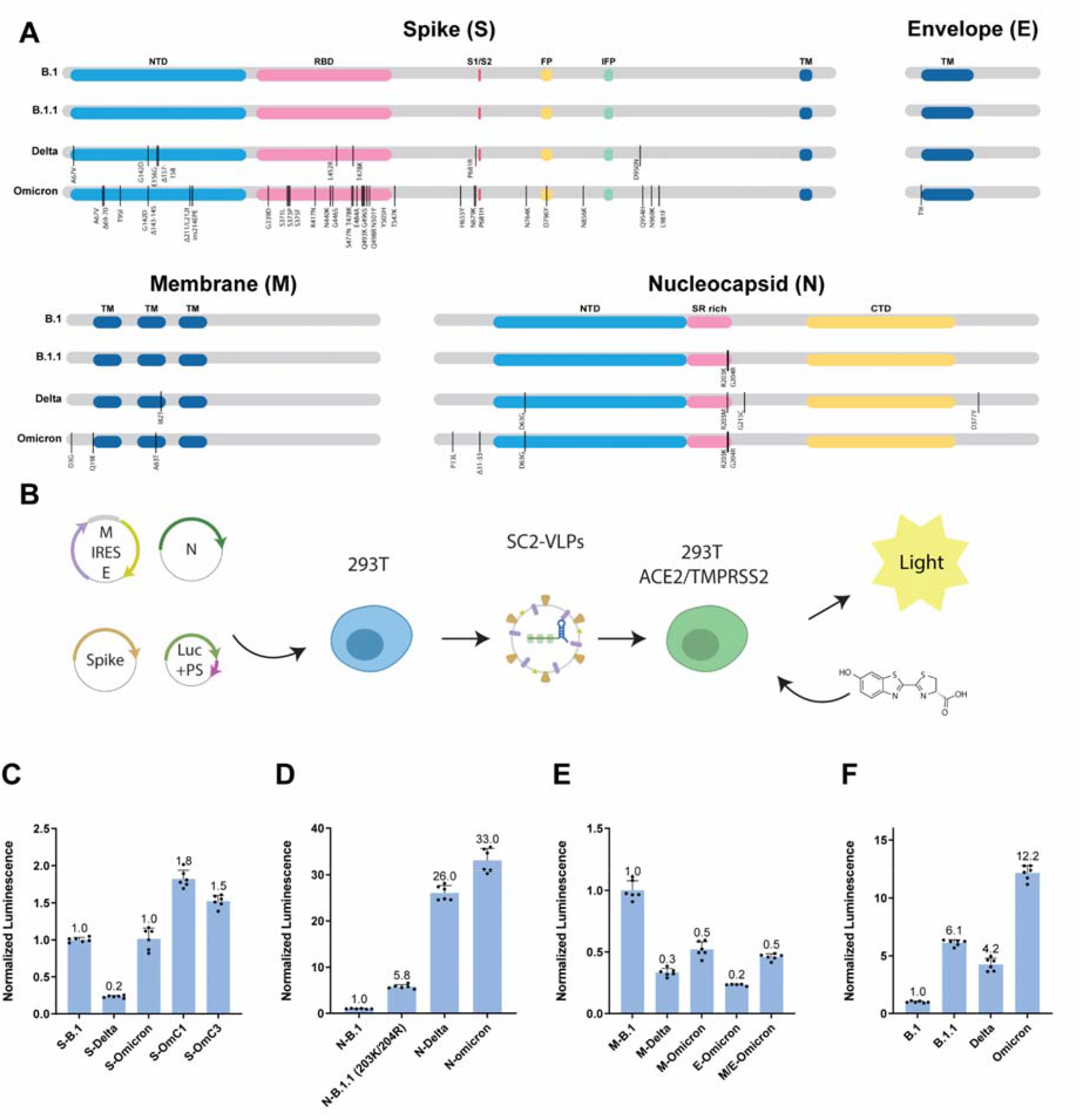
Omicron structural gene variants alter infectivity of SC2-VLPs. (**A**) Sequence differences in genes encoding the structural proteins S, E, M and N between B.1, B.1.1, Delta and Omicron viral variants (vertical lines); Omicron-Class1 and Omicron-Class 3 mutations were created for this study. (**B**) Workflow for generating SC2-VLPs and testing their ability to transduce ACE2- and TMPRSS2-expressing HEK293T cells; SC2-VLPs assembled in packaging cells transformed with plasmids encoding S, E, M and N genes as well as a luciferase mRNA fused to the SARS-CoV-2 packaging signal are tested for receptor-mediated cell transduction using a luciferase detection assay. (**C-F**) Luminescence measured as a function of VLPs generated with the component protein shown, in a background of B.1 genes (see text for details).

We next compared the effects of N, M or E viral variants on infectivity of VLPs generated using B.1 genes. The N gene was found previously to have a pronounced influence on infectivity and RNA packaging efficiency (*3*). Required for replication, RNA binding, packaging, stabilization and release, the N protein includes a seven amino acid mutational hotspot (N:199-205) in a region linking the N- and C-terminal domains. Notably, B.1.1, Delta and Omicron, but not B.1, include mutations at R203 that were found previously to enhance VLP infectivity and RNA packaging (Fig. 1A, Supplementary Table S2) (*3*). The N-Delta and N-Omicron variants generated VLPs with robust infectivity that was enhanced relative to both B.1 and B.1.1 variants, consistent with the N protein playing a central role in viral packaging and cell transduction efficiency (Fig. 1D).

Omicron contains three mutations in the M protein and one mutation in the E protein relative to B.1/Delta. VLPs generated using the Omicron M or E proteins, but with B.1 versions of the other structural components, showed levels of infectivity that were reduced relative to those measured for the B.1 VLPs (Fig. 1E, Supplementary Table S3, 4). This finding suggests that some Omicron mutations reduce viral fitness, at least on their own. To test if these effects are mitigated by mutations in other structural proteins, we also tested VLPs generated using the combined structural protein mutations for each variant. We found that Omicron VLPs were twice as infectious as VLPs generated using Delta or B.1.1 structural proteins and 12-fold more infectious than VLPs generated using B.1 VLPs, consistent with observed enhancements seen in the individual proteins (Fig. 1F).

We next tested the VLP neutralization capability of antisera collected from 38 individuals 4-6 weeks post-vaccination with Pfizer/BioNTech, Moderna or Johnson & Johnson vaccines, or convalescent sera from unvaccinated COVID-19 survivors. The antisera were collected from participants aged 18-50 years enrolled in a clinical trial led by Curative, and SARS-CoV-2 IgG antibodies were quantified with an ELISA (Supplementary Table S5). The serum was heat-inactivated at 56°C for 30 mins and then incubated with VLPs at dilutions 1/20, 1/80, 1/320, 1/1280, 1/5120 and 1/20480 for a total of six dilutions. VLPs were generated with B.1 structural genes except for N-R203M which we previously found to enhance assembly and increase the dynamic range of our neutralization assay. In initial experiments using B.1 spike we found that sera from both Pfizer/BioNTech and Moderna vaccinated individuals yielded high neutralization titers with medians of 549 and 490 respectively (Table 1). Sera from Johnson and Johnson vaccinated and convalescent patients had lower titers with median of 25 and 35 respectively (Table 1) matching the low levels of SARS-CoV-2 IgG antibodies detected in this cohort (Supplementary Table S5).

**Table 1.** Neutralization titers of vaccinated or convalescent individuals against S-variants. Numbers indicate dilution factors that yields 50% neutralization. Higher numbers indicate better neutralization. Red fill indicates undetectable neutralization at the lowest (1/20) dilution.

|  | B.1 | Delta | Omicron | OmC1 | OmC3 |
| --- | --- | --- | --- | --- | --- |
| PF0002 | 5900 | 880 | 768 | 4006 | 2435 |
| PF0004 | 4396 | 1248 | 204 | 1206 | 1244 |
| PF0005 | 549 | 185 | 20 | 172 | 130 |
| PF0006 | 194 | 52 | 17 | 34 | 68 |
| PF0007 | 752 | 319 | 30 | 190 | 357 |
| PF0009 | 1159 | 204 | 178 | 483 | 475 |
| PF0011 | 824 | 241 | 43 | 166 | 289 |
| PF0012 | 282 | 108 | 19 | 57 | 140 |
| PF0013 | 152 | 110 | 18 | 45 | 85 |
| PF0016 | 37 | 1 | 17 | 9 | 31 |
| PF0017 | 295 | 118 | 37 | 110 | 151 |
| M0002 | 3830 | 727 | 692 | 3185 | 1771 |
| M0003 | 375 | 75 | 26 | 102 | 173 |
| M0004 | 25608 | 6105 | 3524 | 15008 | 10995 |
| M0005 | 376 | 130 | 54 | 133 | 174 |
| M0006 | 450 | 80 | 24 | 229 | 178 |
| M0007 | 531 | 131 | 41 | 205 | 215 |
| M0008 | 186 | 76 | 17 | 94 | 111 |
| M0009 | 608 | 168 | 41 | 205 | 245 |
| M0010 | 171 | 35 | 2 | 47 | 60 |
| M0011 | 823 | 158 | 53 | 238 | 232 |
| JJ0002 | 60 | 2 | 16 | 16 | 11 |
| JJ0003 | 58 | 10 | 15 | 6 | 13 |
| JJ0005 | 26 | 7 | 19 | 35 | 15 |
| JJ0006 | 26 | 9 | 16 | 10 | 13 |
| JJ0007 | 11 | 12 | 14 | 7 | 18 |
| JJ0008 | 25 | 16 | 55 | 14 | 19 |
| JJ0009 | 10 | 8 | 14 | 0 | 14 |
| JJ0010 | 15 | 7 | 20 | 6 | 7 |
| JJ0011 | 20 | 5 | 12 | 3 | 12 |
| PC0002 | 51 | 44 | 43 | 19 | 12 |
| PC0003 | 7 | 22 | 20 | 9 | 25 |
| PC0006 | 5 | 0 | 15 | 5 | 5 |
| PC0007 | 31 | 0 | 24 | 12 | 24 |
| PC0008 | 39 | 323 | 27 | 14 | 26 |
| PC0009 | 268 | 113 | 24 | 104 | 14 |
| PC0011 | 432 | 19044 | 77 | 44 | 291 |
| PC0013 | 0 | 112 | 8 | 0 | 30 |
| Naïve | 5 | 11 | 19 | 9 | 2 |

We then substituted the S-gene with S-variants previously tested, as they have varying mutations in the RBD known to affect neutralization. We tested the neutralization capacity of each patient’s serum against VLPs representing S from B.1, Delta or Omicron viral variants (Fig. 2A-D). There was a pronounced decrease of 15- to 18-fold in potency against Omicron, with intermediate effects on Delta, for the antisera from mRNA vaccine recipients (Fig. 2A-D; Table 1). Limited efficacy was detected for any of the sera from those vaccinated with the adenovirus based Johnson and Johnson vaccine and variable neutralization was observed for COVID-19 survivors (Table 1). We next examined whether Class 1 or Class 3 mutations were responsible for reduced neutralization in patient anti sera. For both S-OmC1 and OmC3 cases we found intermediate neutralization, suggesting that neutralization escape from patient sera is a function of several mutations acting in concert (Supplementary Fig. S1A-D). Third-dose vaccination with the Pfizer vaccine increased titers against all variants including Omicron (Fig. 2E-H, Supplementary Fig. S1E-G, Table 2) as measured at 16 and 21 days after the third dose. All 8 sera from this cohort had low (median 64) neutralization titers against Omicron 21 days after their third dose while only 1 out 8 had detectable neutralization prior to boosting (Fig. 2G). However, even after boosting we observed 8-fold reduced neutralizing titers against Omicron compared to B.1, suggesting that Omicron is still able to partially escape neutralizing antibodies induced by vaccination with the ancestral S-B.1 (Fig. 2H, Table 2).

**Table 2:** Neutralization titers against S-variants of individuals vaccinated with two or three doses of the Pfizer vaccine. Each row represents one subject. Numbers indicate dilution factors that yield 50% neutralization. Higher numbers indicate better neutralization. Red fill indicates undetectable neutralization at the lowest (1/20) dilution. Last three columns indicate the time elapsed between doses for each individual.

| NT50 against S-B.1 |  |  | NT50 against S-Delta |  |  | NT50 against S-Omicron |  |  | Time Lapsed between samples |  |  |
| --- | --- | --- | --- | --- | --- | --- | --- | --- | --- | --- | --- |
| Pre boost | T1 | T2 | Pre boost | T1t | T2 | Pre boost | T1 | T2 | T0 (Third dose-Second Dose) days | T1 (Days post booster shot) | T2 (Days post booster shot) |
| 9 | 222 | 238 | 2 | 60 | 55 | 0 | 55 | 54 | 239 | 14 | 20 |
| 977 | 2251 | 2070 | 254 | 664 | 593 | 58 | 135 | 126 | 194 | 17 | 21 |
| 120 | 3311 | 3213 | 31 | 816 | 631 | 5 | 512 | 474 | 215 | 17 | 20 |
| 0 | 139 | 274 | 0 | 32 | 84 | 1 | 27 | 58 | 197 | 19 | 22 |
| 13 | 473 | 378 | 0 | 138 | 127 | 2 | 52 | 53 | 190 | 16 | 21 |
| 3 | 448 | 432 | 0 | 124 | 116 | 0 | 47 | 46 | 212 | 17 | 20 |
| 57 | 1537 | 1197 | 6 | 444 | 404 | 3 | 260 | 274 | 200 | 17 | 20 |
| 19 | 404 | 477 | 11 | 130 | 147 | 0 | 46 | 69 | 239 | 17 | 22 |

**Fig. 2.**
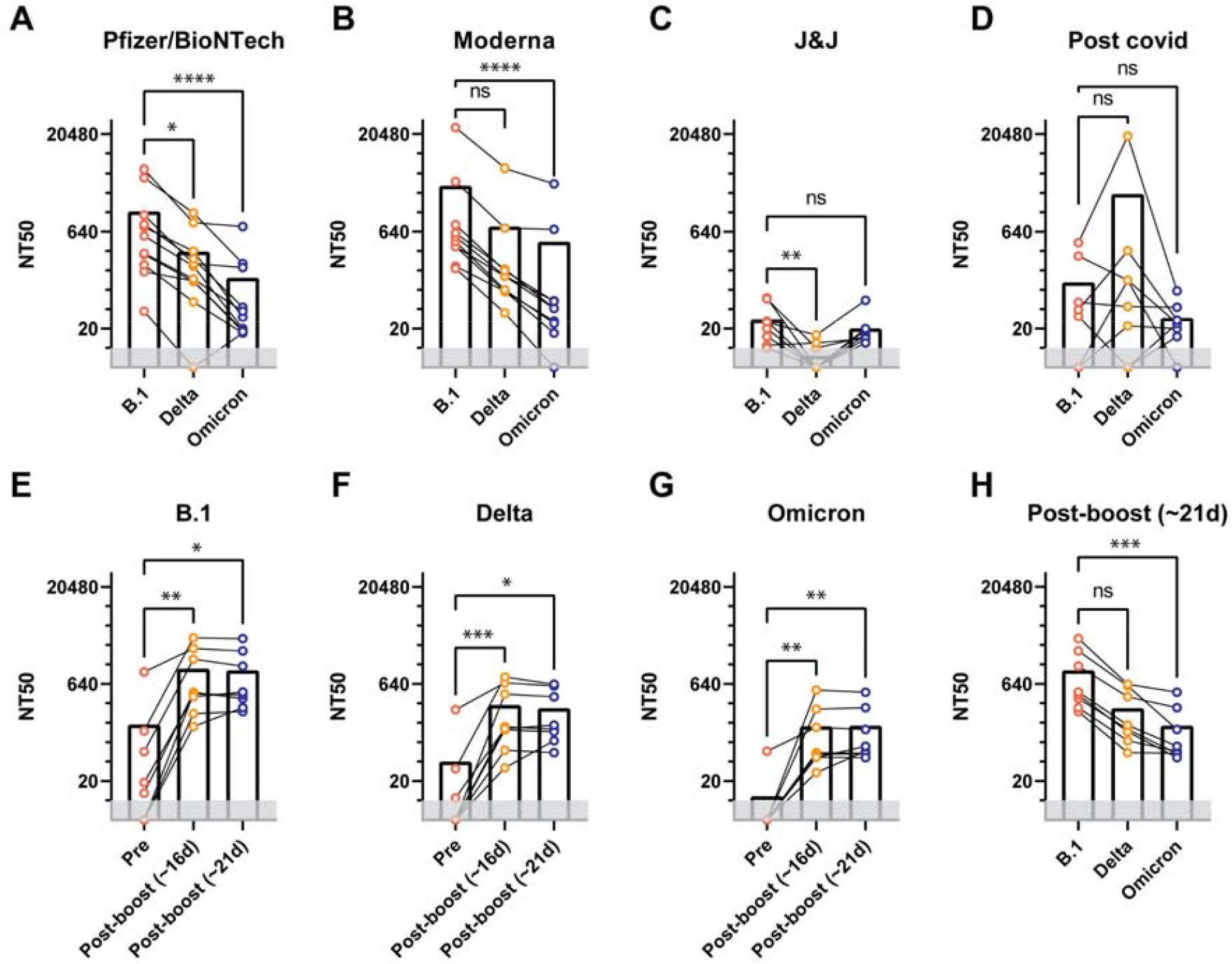
Antiserum neutralization of VLPs generated with different S genes. (A-D) 50% neutralization titers of sera isolated from individuals vaccinated using Pfizer/BioNTech, Moderna, Johnson and Johnson vaccines or from convalescent COVID-19 patients. Neutralization curves were determined using VLPs with either S-B.1, S-Delta, S-Omicron, S-OmC1 or S-OmC3. (E-H) Neutralization titers of sera collected before and after third dose vaccination from individuals receiving the Pfizer/BioNTech vaccine. *p<0.05, **p<0.01, ***p<0.001, ****p<0.0001 evaluated using Friedman’s exact test for repeated measures.

To test whether monoclonal antibodies generated against the ancestral SARS-CoV-2 S protein would be effective at Omicron neutralization, VLPs were generated using the Omicron, OmC1 or OmC3 S genes, and transduction assays were conducted in the presence or absence of Class 1 (Casirivimab) or Class 3 (Imdevimab) monoclonal antibodies (Fig. 3A-E, Table 3). Strikingly, although both antibodies had robust neutralization activity against B.1.1 or Delta VLPs, no activity was detected for either one against Omicron VLPs. When the OmC1 or OmC3 versions of the S gene were tested in this VLP assay, we found that Casirivimab was able to neutralize OmC3 but not OmC1 and Imdevimab was able to neutralize OmC1 but not OmC3. This suggests that the six mutations within the Omicron RBD (K417N, N440K, G446S, G496S, Q498R, N501Y) are largely responsible for the failure of these monoclonal antibodies to neutralize Omicron S.

**Table 3:**
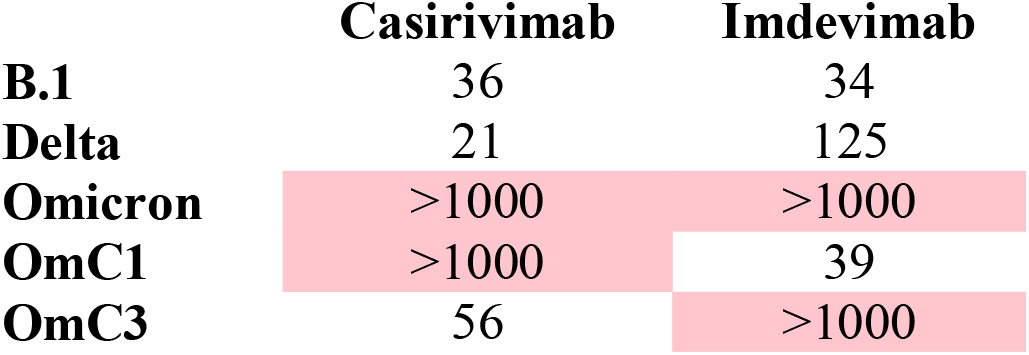
IC50 of Casirivimab and Imdevimab against S variants (ng/mL). Smaller numbers indicate better neutralization. Red fill indicates undetectable neutralization in our assay of >1000ng/mL.

**Fig. 3.**
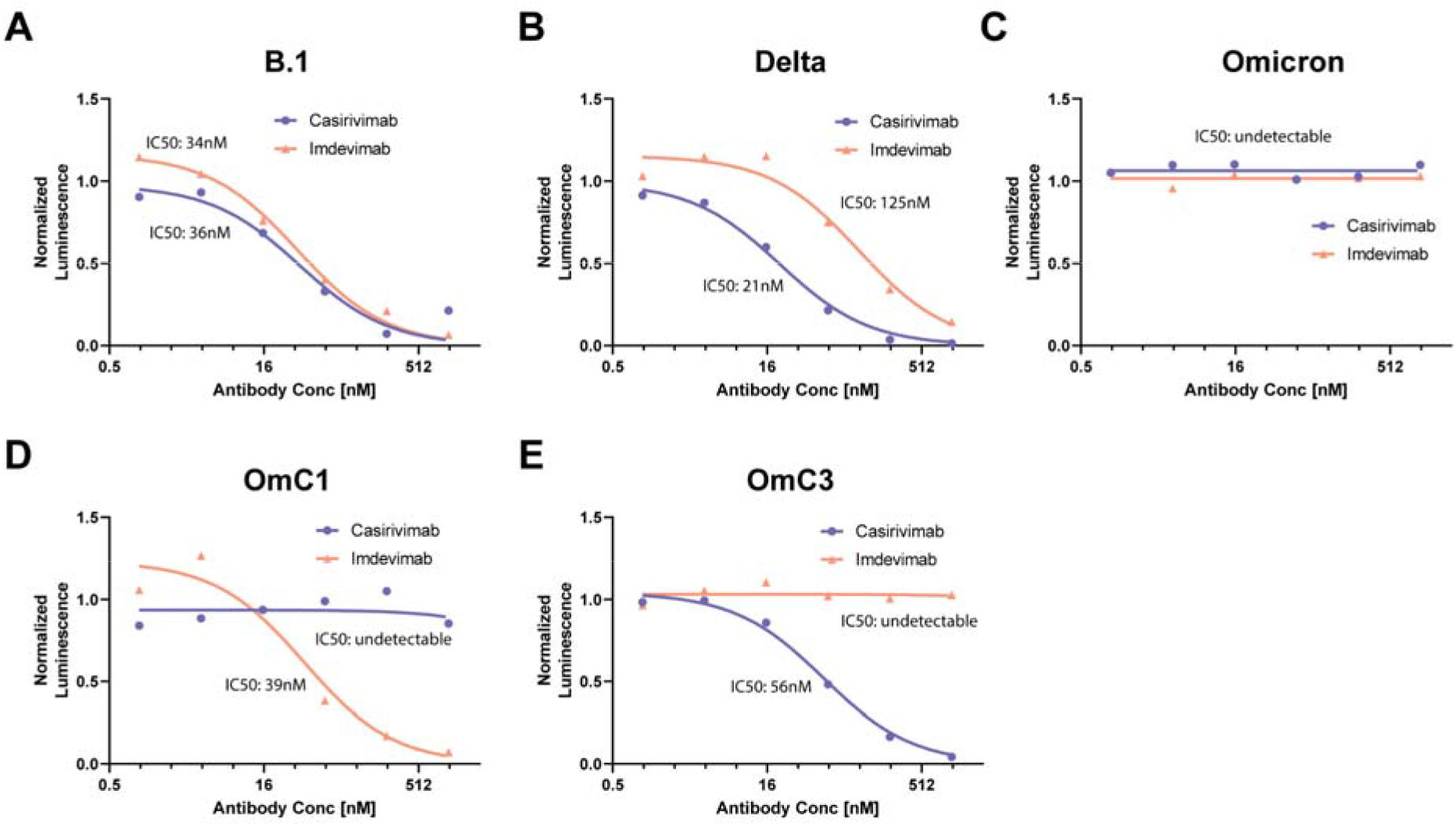
Antibody neutralization of VLPs generated with different S genes. Neutralization curves and IC50 values of Casirivimab and Imdevimab against the S-variants: S-B.1, S-Delta, S-Omicron, S-OmC1 or S-OmC3. Inset shows IC50 values for each monoclonal antibody.

In summary, SARS-CoV-2 virus-like particles that transduce reporter mRNA into ACE2- and TMPRSS2-expressing cells enabled a rapid and comprehensive comparison of structural protein (S, E, M, N) variant effects on both particle infectivity and antibody neutralization. Using this system, we found that the Omicron versions of both S and N enhance VLP infectivity relative to ancestral viral variants including Delta. Omicron maintains mutations in the N mutational hotspot that were shown previously to confer markedly enhanced VLP infectivity (*3*). Surprisingly, Omicron M and E gene variants appear to compromise infectivity, at least in the context of ancestral versions of the other structural genes, implying that genes including S and N override less fit versions of M, E and perhaps other genes in the intact virus. Monitoring S and N gene evolution and determining why the N gene has such a pronounced effect on viral particle infectivity may enable development of better diagnostics, more broadly neutralizing vaccine development and potentially new therapeutics.

Notably, all antisera from vaccinated individuals or convalescent sera from COVID-19 survivors showed reduced neutralization of Omicron VLPs relative to ancestral variants including Delta, with mRNA vaccines far surpassing a viral vector vaccine or natural infection in initial potency. These data do not account for T cell-based immunity induced by vaccination or prior infection. We also found that Omicron S mutations interfere with Class 1 and Class 3 monoclonal antibody binding, rendering some commercially available therapeutic antibodies completely ineffective. These results suggest that prior to vaccine boost, antibodies produced by mRNA vaccines have 15- to 18-fold reduced efficacy against Omicron, and that the Johnson and Johnson vaccine produces limited neutralizing antibodies against any SARS-CoV-2 variant. Booster shots increase neutralization titers against Omicron but the titers remain much lower than for previous variants. Consistent with data from recent pseudovirus neutralization studies (*5, 6*), these findings support the use of mRNA vaccine boosters to enhance antibody-based protection against Omicron infection, in lieu of vaccines tailored to Omicron itself.

## Data Availability

All data produced in the present work are contained in the manuscript.

## Acknowledgments

We thank members of the Doudna and Ott labs for helpful discussions.

## Funding

This project was funded by a grant to JAD from the National Institutes of Health (R21AI59666) and by support from the Howard Hughes Medical Institute and the Gladstone Institutes. AMS acknowledges support from the National Sciences and Engineering research Council of Canada (NSERC PDF-533021-2019), IPC received support from the NIH (F31AI164671-01), and MO received support from the Roddenberry Foundation and a gift from Pam and Ed Taft.

## Author contributions

Conceptualization: AMS, GRK, MO, JAD

Anti sera: IS, NK, BM, VH, MS, LL, MB, FT, LS

Investigation: AMS, AC, MMK, BS Funding acquisition: MO, JAD

Project administration/coordination: GRK

Supervision: MO, JAD

Writing – original draft: AMS, JAD

Writing – review & editing: AMS, JAD, LS, GRK, MO

## Competing interests

AMS and JAD are inventors on a patent application filed by the Gladstone Institutes and the University of California that covers the method and composition of SARS-CoV-2 VLP preparations for RNA transduction and expression in cells.

## Data and materials availability

All data are available in the main paper or supplementary material. Plasmids are available from Addgene (addgene.org) or by request. All other materials available upon request.

## Supplementary Materials

Materials and Methods

Supplementary Text

Figs. S1

Tables S1 to S5

References (*##*–*##*)

## Notes

### Author Declarations

Ethics committee/IRB of Advarra gave ethical approval for this work

### Summary of Updates

This version has been revised to fix a mistake in the units in Figure 3. Changed from nM to ng/mL.

## References and Notes

1. K. H. D. Crawford, R. Eguia, A. S. Dingens, A. N. Loes, K. D. Malone, C. R. Wolf, H. Y. Chu, M. A. Tortorici, D. Veesler, M. Murphy, D. Pettie, N. P. King, A. B. Balazs, J. D. Bloom, Protocol and Reagents for Pseudotyping Lentiviral Particles with SARS-CoV-2 Spike Protein for Neutralization Assays. Viruses. 12 (2020), doi:10.3390/v12050513.

2. J. A. Plante, Y. Liu, J. Liu, H. Xia, B. A. Johnson, K. G. Lokugamage, X. Zhang, A. E. Muruato, J. Zou, C. R. Fontes-Garfias, D. Mirchandani, D. Scharton, J. P. Bilello, Z. Ku, Z. An, B. Kalveram, A. N. Freiberg, V. D. Menachery, X. Xie, K. S. Plante, S. C. Weaver, P.-Y. Shi, Spike mutation D614G alters SARS-CoV-2 fitness. Nature. 592, 116–121 (2021).

3. A. M. Syed, T. Y. Taha, T. Tabata, I. P. Chen, A. Ciling, M. M. Khalid, B. Sreekumar, P.-Y. Chen, J. M. Hayashi, K. M. Soczek, M. Ott, J. A. Doudna, Rapid assessment of SARS-CoV-2 evolved variants using virus-like particles. Science, eabl6184 (2021).

4. A. J. Greaney, T. N. Starr, P. Gilchuk, S. J. Zost, E. Binshtein, A. N. Loes, S. K. Hilton, J. Huddleston, R. Eguia, K. H. D. Crawford, A. S. Dingens, R. S. Nargi, R. E. Sutton, N. Suryadevara, P. W. Rothlauf, Z. Liu, S. P. J. Whelan, R. H. Carnahan, J. E. Crowe Jr, J. D. Bloom, Complete Mapping of Mutations to the SARS-CoV-2 Spike Receptor-Binding Domain that Escape Antibody Recognition. Cell Host Microbe. 29, 44–57.e9 (2021).

5. S. Cele, L. Jackson, K. Khan, D. S. Khoury, T. Moyo-Gwete, H. Tegally, C. Scheepers, D. Amoako, F. Karim, M. Bernstein, G. Lustig, D. Archary, M. Smith, Y. Ganga, Z. Jule, K. Reedoy, D. Cromer, J. E. San, S.-H. Hwa, J. Giandhari, J. M. Blackburn, B. I. Gosnell, S. S. A. Karim, W. Hanekom, NGS-SA, COMMIT-KZN Team, A. von Gottberg, J. Bhiman, R. J. Lessells, M.-Y. S. Moosa, M. P. Davenport, T. de Oliveira, P. L. Moore, A. Sigal, SARS-CoV-2 Omicron has extensive but incomplete escape of Pfizer BNT162b2 elicited neutralization and requires ACE2 for infection. medRxiv (2021), doi:10.1101/2021.12.08.21267417.

6. L. Lu, B. W.-Y. Mok, L.-L. Chen, J. M.-C. Chan, O. T.-Y. Tsang, B. H.-S. Lam, V. W.-M. Chuang, A. W.-H. Chu, W.-M. Chan, J. D. Ip, B. P.-C. Chan, R. Zhang, C. C.-Y. Yip, V. C.-C. Cheng, K.-H. Chan, D.-Y. Jin, I. F.-N. Hung, K.-Y. Yuen, H. Chen, K. K.-W. To, Neutralization of SARS-CoV-2 Omicron variant by sera from BNT162b2 or Coronavac vaccine recipients. Clin. Infect. Dis. (2021), doi:10.1093/cid/ciab1041.

