## Supplemental Information for "Omicron mutations enhance infectivity and reduce antibody neutralization of SARS-CoV-2 virus-like particles"

**This PDF file includes:**

Materials and Methods
Supplementary Text
Figs. S1
Tables S1 to S5

### Materials and Methods

*Cloning for plasmids encoding structural proteins:* pcDNA3.1 backbone plasmids were generated encoding N, and M-IRES-E. Sequences for E, M and N were PCR amplified from codon optimized plasmids were gifts from Nevan Krogan (Addgene plasmid # 141385, 141386, 141391, ). pcDNA3.1-SARS2-Spike was a gift from Fang Li (Addgene plasmid # 145032). Site directed mutagenesis (NEB) was used to remove the C9-tag and introduce the D614G mutation. Delta and Omicron structural protein were cloned ligating eBlocks (IDT) gene fragments following NEBuilder HiFi DNA (NEB E2621L) Assembly Reaction Protocol.

*SC2-VLP production:* For a 24-well, plasmids CoV2-N (0.67), CoV2-M-IRES-E (0.33), CoV-2-Spike (0.006) and Luc-PS9 (1.0) at indicated mass ratios for a total of 1 µg of DNA were diluted in 50 µL Opti-MEM. 3 µg PEI was diluted in 50 µL Opti-MEM and added to plasmid dilution quickly to complex the DNA. Transfection mixture was incubated for 20 minutes at room temperature and then added dropwise to 293T cells in 0.5 mL of DMEM containing fetal bovine serum and penicillin/streptomycin. Media was changed after 24 hours of transfection and At 48 hours post-transfection, VLP containing supernatant was collected and filtered using a 0.45 µm syringe filter. For other culture sizes, the mass of DNA used was 1 µg for 24-well, 4 µg for 6-well, 20 µg for 10-cm plate and 40 µg for 15-cm plate. Opti-MEM volumes were 100 µL, 400 µL, 1 mL and 3 mL respectively and PEI was always used at 3:1 mass ratio.

*Luciferase readout*: In each well of a clear 96-well plate 50 µL of SC2-VLP containing supernatant was added to 50 µL of cell suspension containing 50 000 receiver cells (293T ACE2/TMPRSS2). Cells were allowed to attach and take up VLPs overnight. Next day, supernatant was removed and cells were lysed in 20 µL passive lysis buffer (Promega) for 15 minutes at room temperature with gentle rocking. Lysates were transferred to an opaque white 96-well plate and 30 µL of reconstituted luciferase assay buffer was added and mixed with each lysate. Luminescence was measured immediately after mixing using a TECAN plate reader with no attenuation and a luminescence integration time of 1 s.

*Western blot and Imaging*: Cells were lysed in RIPA buffer (Thermo Scientific™ 89900) and lysates were collected after centrifugation. Laemmli loading buffer (1x final) and dithiothreitol (DTT, 40 mM final) was added to lysates and heated for 95°C for 5 minutes to lyse denature proteins. Samples were loaded onto 4-20% gradient gels (Biorad 567-1094) and transferred to a PVDF membrane (Biorad). Membrane was blocked in 10% NFDM and stained with primary antibody: anti-N (abcam ab273434, 1:500 dilution), anti-S (abcam ab272504, 1:1000), anti-GAPDH (Cell signaling D16H11), 1:5000) for 4 hours at room temperature or O/N at 4°C. Blots were rinsed with TBS-T three times for 10 minutes each and stained with secondary (abcam ab205719 (mouse), 1:5000; Bethyl A120-201P(Rabbit)). Imaged using pierce chemiluminescence kit.

*Cell lines:* Cells were maintained in a humidified incubator at 37°C in 5% CO2 in the indicated media and passaged every 3-4 days. 293T cells were obtained from ATCC and maintained in DMEM with 10% FBS and 1% penicillin/streptomycin.. 293T cells stably co-expressing ACE2 and TMPRSS2 were generated through sequential transduction of 293T cells with TMPRSS2-encoding (generated using Addgene plasmid #170390, a gift from Nir Hacohen and ACE2-encoding (generated using Addgene plasmid #154981, a gift from Sonja Best) lentiviruses and selection with hygromycin (250 µg/mL) and blasticidin (10 µg/mL) for 10 days, respectively. ACE2 and TMPRSS2 expression was verified by western blot.

*Neutralization Assay:* Each heat inactivated serum sample was serially diluted from 1:20 to 1:20480 dilution in complete DMEM media prior to incubation (1hr at 37°C) with 40μL VLP with total volume of 50μL, then plated onto receiver cells (50000 293T ACE2-TMPRSS2 cells). 24hr later luciferase readout was taken. Neutralization (NT50) was estimated by interpolating the dilution of serum at which 50% infectivity was reduced.

*Serum samples:* Serum samples from individuals not exposed to SARS-CoV-2 (pre-COVID, control), exposed to SARS-CoV-2 (post-COVID), and those vaccinated with either two doses of elasomeran (Moderna), two doses of tozinameran (Pfizer/BioNTech) vaccine or one dose of Johnson & Johnson vaccine were collected through a clinical trial led by Curative (Supplementary Table 5). Post-COVID samples reflect non vaccinated participant samples that were collected within 4-6 weeks of the original positive test and were negative by PCR at the time of serum collection. Serum from vaccinated participants was collected 4-6 weeks post vaccination following final dose. The clinical trial protocol was approved by Advarra under Pro00054108 for a study designed to investigate immune escape by SARS-CoV-2 variants. The trial has been submitted to clinicaltrials.gov registry (NCT ID pending, Unique Protocol ID: PTL-2021-0007). Sample specimens were collected from adult individuals aged 18 to 50 years who either had been vaccinated for COVID-19 and/or had a history of COVID-19. Vulnerable populations were excluded from enrollment. Patients signed consent forms held by Curative. Participants were enrolled from individuals that tested with Curative in Los Angeles County and were sent an IRB-approved email enrollment script. Those who were interested were contacted by the Curative Clinical Trials research team (CITI trained) and those who consented to the study were scheduled for sample collection by a clinician who went to their residence. Participants underwent a standard venipuncture procedure. Briefly, licensed phlebotomists collected a maximum of 15 ml whole blood. Once collected, the sample was left at ambient temperature for 30–60 min to coagulate, then was centrifuged at 2200–2500 rpm for 15 min at room temperature. Samples were then placed on ice until delivered to the laboratory site where the serum was aliquoted to appropriate volumes for storage at −80 ºC until use. A quantitative SARS-CoV-2 IgG ELISA was performed on serum specimens (EuroImmun, Anti-SARS-CoV-2 ELISA (IgG), 2606–9621G, New Jersey). To quantify SARS-CoV-2 IgG antibodies, an S1-specific monoclonal IgG antibody with no known cross-reactivity to the S2 domain of the spike protein was used as a reference antibody. A standard curve was developed using a monoclonal IgG antibody targeting the S1 antigen of SARS-CoV-2 at different concentrations with a polynomial regression curve-fitting model. The standard curve was used to calculate the sample IgG antibody concentration. Serum samples were heat inactivated at 56°C for 30 mins prior to use in VLP assays. Pre-COVID sera was pooled into one sample.

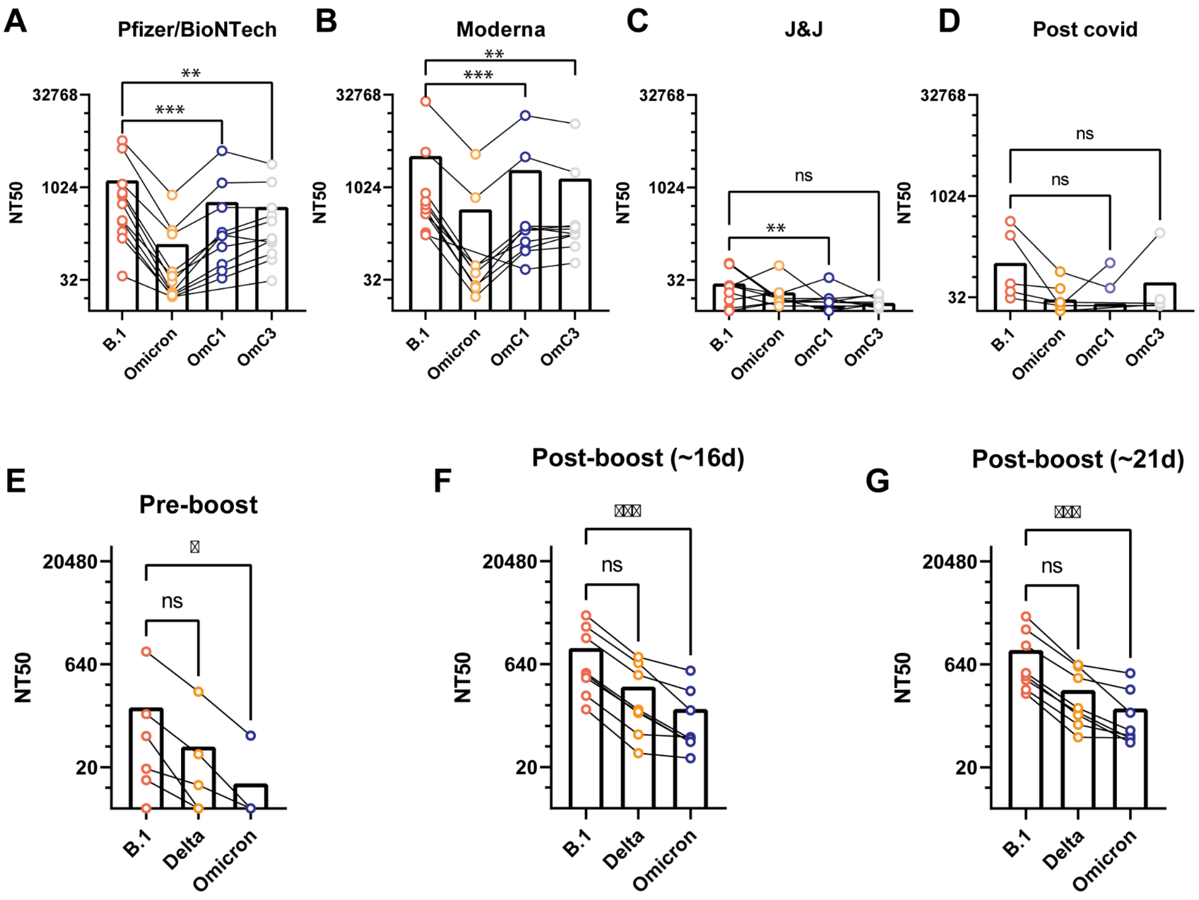


**Fig. S1. Neutralization titers of subjects vaccinated and boosted against SARS-CoV-2.** (A-D) Neutralization titers of subjects vaccinated with Pfizer/BioNTech, Moderna, or J&J or subjects recovered from infection. Neutralization evaluated against the ancestral B.1 lineage S protein, S-Omicron or a modified S-Omicron containing mutations targeting Class 1 and Class 3 antibodies targeting the receptor binding domain. (E-G) Effect of third dose booster for subjects vaccinated using the Pfizer/BioNTech vaccine evaluated against B.1, Delta or Omicron at ~16 days post booster or 21 days post booster.

A

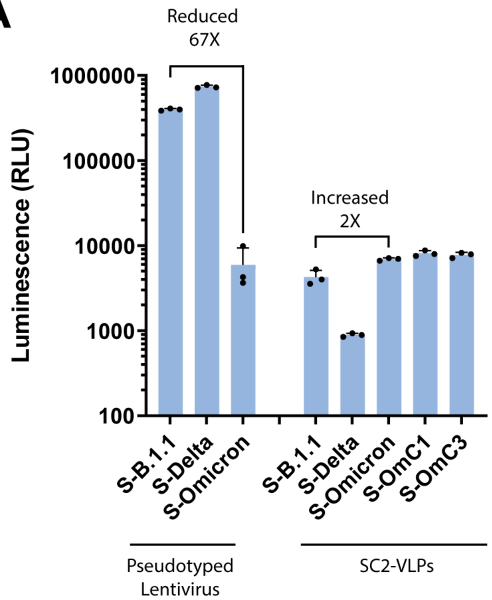


**Fig. S2. Comparison of S-variants using pseudotyped lentivirus and VLPs.** Luciferase signal from cells transduced using S-pseudotyped lentiviruses or VLPs. Delta and Omicron have opposite trends in infectivity in these two systems.

**Table S1.**
List of mutations used in each S variant divided by domains.

| | NTD | RBD | CTD |
|---|---|---|---|
| **B.1** | | | D614G |
| **B.1.1** | | | D614G |
| **Delta** | A67V, G142D, E156G, Δ157-158 | L452R, T478K | D614G, P681R, D950N |
| **Omicron** | A67V, Δ69-70, T95I, G142D, Δ143-145, Δ211, L212I, ins214-EPE | G339D, S371L, S373P, S375F, K417N, N440K, G446S, S477N, T478K, E484A, Q493K, G496S, Q498R, N501Y, Y505H | T547K, D614G, H655Y, N679K, P681H, N764K, D796Y, N856K, Q954H, N969K, L981F |
| **OmC1** | A67V, Δ69-70, T95I, G142D, Δ143-145, Δ211, L212I, ins214-EPE | K417N, G496S, Q498R, N501Y | T547K, D614G, H655Y, N679K, P681H, N764K, D796Y, N856K, Q954H, N969K, L981F |
| **OmC3** | A67V, Δ69-70, T95I, G142D, Δ143-145, Δ211, L212I, ins214-EPE | N440K, G446S, G496S, Q498R | T547K, D614G, H655Y, N679K, P681H, N764K, D796Y, N856K, Q954H, N969K, L981F |

**Table S2.**
List of mutations used in each N variant divided by domains.

| | NTD | SR | linker | CTD |
|---|---|---|---|---|
| **B.1** | | | | |
| **B.1.1** | | R203K, G204R | | |
| **Delta** | D63G | R203M | G215C | D377Y |
| **Omicron** | P13L, Δ31-33, D63G | R203K, G204R | | |

**Table S3.**
List of mutations used in each M variant.

| | |
|---|---|
| **B.1** | |
| **B.1.1** | |
| **Delta** | I82T |
| **Omicron** | D3G, Q19E, A63T |

**Table S4.**
List of mutations used in each E variant.

| | |
|---|---|
| **B.1** | |
| **B.1.1** | |
| **Delta** | |
| **Omicron** | T9I |

**Table S5.**

Serum samples from clinical trial participants used in VLP assays in this study.

| Subject ID | ELISA Status | Total IgG (ug/ml) | Sample Type |
|---|---|---|---|
| CUR01 | Negative | / | pre-COVID serum |
| CUR02 | Negative | / | pre-COVID serum |
| CUR03 | Negative | / | pre-COVID serum |
| CUR04 | Negative | / | pre-COVID serum |
| CUR05 | Negative | / | pre-COVID serum |
| PC0002 | Positive | 4.45 | post-COVID serum |
| PC0003 | Positive | 0.44 | post-COVID serum |
| PC0006 | Positive | 2.29 | post-COVID serum |
| PC0007 | Positive | 1.19 | post-COVID serum |
| PC0008 | Positive | 2.16 | post-COVID serum |
| PC0009 | Positive | 1.19 | post-COVID serum |
| PC0011 | Positive | 39.8 | post-COVID serum |
| PC0013 | Positive | 1.03 | post-COVID serum |
| PF0002 | Positive | 9.67 | Pfizer vaccinee serum - 2 doses |
| PF0004 | Positive | 9.32 | Pfizer vaccinee serum - 2 doses |
| PF0005 | Positive | 9.36 | Pfizer vaccinee serum - 2 doses |
| PF0006 | Positive | 5.05 | Pfizer vaccinee serum - 2 doses |
| PF0007 | Positive | 8.85 | Pfizer vaccinee serum - 2 doses |
| PF0009 | Positive | 8.21 | Pfizer vaccinee serum - 2 doses |
| PF0011 | Positive | 9.66 | Pfizer vaccinee serum - 2 doses |
| PF0012 | Positive | 7.01 | Pfizer vaccinee serum - 2 doses |
| PF0013 | Positive | 6.41 | Pfizer vaccinee serum - 2 doses |
| PF0016 | Positive | 1.79 | Pfizer vaccinee serum - 2 doses |
| PF0017 | Positive | 7.72 | Pfizer vaccinee serum - 2 doses |
| M0002 | Positive | 91.77 | Moderna vaccinee serum - 2 doses |
| M0003 | Positive | 14.5 | Moderna vaccinee serum - 2 doses |
| M0004 | Positive | 71.94 | Moderna vaccinee serum - 2 doses |
| M0005 | Positive | 9.88 | Moderna vaccinee serum - 2 doses |
| M0006 | Positive | 8.5 | Moderna vaccinee serum - 2 doses |
| M0007 | Positive | 10.5 | Moderna vaccinee serum - 2 doses |
| M0008 | Positive | 21.38 | Moderna vaccinee serum - 2 doses |
| M0009 | Positive | 10.2 | Moderna vaccinee serum - 2 doses |
| M0010 | Positive | 15.65 | Moderna vaccinee serum - 2 doses |

| | | | |
|---|---|---|---|
| M0011 | Positive | 15.08 | Moderna vaccinee serum - 2 doses |
| JJ0002 | Positive | 1.09 | J+J vaccinee serum - 1 dose |
| JJ0003 | Positive | 1.63 | J+J vaccinee serum - 1 dose |
| JJ0005 | Positive | 1.29 | J+J vaccinee serum - 1 dose |
| JJ0006 | Positive | 2.09 | J+J vaccinee serum - 1 dose |
| JJ0007 | Positive | 1.19 | J+J vaccinee serum - 1 dose |
| JJ0008 | Positive | 1.84 | J+J vaccinee serum - 1 dose |
| JJ0009 | Positive | 0.57 | J+J vaccinee serum - 1 dose |
| JJ0010 | Positive | 0.55 | J+J vaccinee serum - 1 dose |
| JJ0011 | Positive | 1.68 | J+J vaccinee serum - 1 dose |
